# How to Develope a Novel 3D Printed Integral Customized Anatomical Acetabular Prosthesis in Hip Arthroplasty for Crowe Type II and III Developmental Dysplasia of the Hip?

**DOI:** 10.1101/2022.11.28.22282849

**Authors:** Heng Zhang, Xiaodong Ma, Zengjing Cheng, Xuanxuan Li, Jiansheng Zhou, Jianning Zhao

## Abstract

**Background:** The acetabular pathomorphology in patients with Crowe type II and III developmental dysplasia of the hip(DDH)often present complicated changes, which bring challenges to the anatomical reconstruction of acetabulum in total hip arthroplasty (THA). The objective of this study was to develope a novel 3D printed integral customized acetabular prosthesis, which provides a promising way to reconstruct the acetabulum with higher accuracy and efficiency by digital softwares, compared with previous 3D printing model method.

**Methods:** 15 patients (18 hips) with end-stage hip osteoarthritis due to Crowe type II/III DDH who underwent primary cementless THA from January 2015 to November 2021 were included. The 15 patients consisted of 3 men (3 hips) and 12 women (15 hips) with an average age of 53.89 ± 8.48 years (range from 32 to 69 years). The novel 3D printed integral customized acetabular prosthesis was designed by the model method and software method respectively. The indicators of the cup size, the volume and superficial area of bone defect, the inclination and anteversion of acetabular cup, the horizontal and vertical distance of hip center and working time were compared between two methods.

**Results:** There were statistically significant difference between the software group and model group on the volume and superficial area of bone defect as well as working time (t = 2.397, 2.707,138.509, P < 0.05). The mean anteversion and inclination of acetabular cup in the software group and model group were(15.17±0.52)°,(40.24±0.58)°and (33.79 ±13.43)°, (30.50 ±11.03)°respectively, the difference was statistically significant (t = 5.859, 3.767, P < 0.05).There were no statistically significant difference between the software group and model group on cup size, the horizontal and vertical distance of hip center(t =1.458,0.114, 1.712, P > 0.05)

**Conclusions:** The application of digital softwares could design and develope the novel 3D printed integral customized anatomical acetabular prosthesis in THA for □Crowe type II and III DDH with high accuracy and efficiency.

## Introduction

The morphology of acetabulum in Crowe type II and III developmental dysplasia of the hip(DDH)present to be disc-shaped and shell-shaped, shallow socket, increased anteversion and inclination, a large number of osteophytes proliferate and segmental bone defects in the superior and posterior parts (1-3). These changes pose challenges for anatomical installation of acetabular prosthesis, precise reconstruction of bone defects and realization of good initial stability of the implants in total hip arthroplasty (THA) (4-6). How to realize anatomical reconstruction of the acetabulum in patients with Crowe type II/III DDH duirng THA and avoid postoperative hip instability, dislocation, lower limb length discrepancy, as well as reduce wear rate and recover hip function are difficult problems confronted by orthopedic surgeons.

It has been accepted that high placement of hip rotation center could simply the operation, increase the coverage rate of acetabular prosthesis, and improve the initial stability of the acetabular cup. However, it is impossible to avoid the improvement of biological stress of the hip joint, the increase of wear rate of the prosthesis, the secondary complications of lower limb length discrepancy, impingement, gluteus medius fatigue, gait changes, dislocation due to the upward migration of the hip rotation center(7-9). Currently, more and more scholars (6, 10, 11) proposed anatomical restoration of rotation center in THA for patients with Crowe type II/III DDH, which is beneficial to balance the stress of ligaments and muscles surrounding the hip joint, reduce the wear rate and loosening of the hip prosthesis, whereas it is difficult to restore the hip rotation center and reconstruct the bone defect during the operation.

Based on previous studies of restoring hip rotation center by locating the acetabular center using the acetabular fossa and acetabular notches as the anatomical landmarks, we developed the 3D printed integral customized acetabular prosthesis, which realized the restoration of hip rotation center and the reconstruction of bone defect simultaneously(12). This method avoided the complications such as unbalanced stress distribution of the joint and soft tissues, the increase of wear rate and loosening of the prosthesis, lower limb length discrepancy, bone resorption, collapse and long-term loosening of the prosthesis caused by structural bone graft in the techniques of high placement of hip center.

In our previous developing process of 3D printed integral customized acetabular prosthesis, as shown in Figure 1, we firstly printed the pelvis model by a 3D printing machine (Arigin 3DM400, Shanghai Arigin Medical Technology Co. LTD, China) using polylactic acid (PLA) material (Fig. 1A). The morphology of acetabular fossa was restored by removing its superficial osteophytes (Fig. 1B). The acetabular center was located at mean 28.7mm point (range, 25–31mm, depending on acetabulum size) above the vertical bisection of the line between the anterior and posterior acetabular notches(3) (Fig.1C). The acetabular socket was reamed by regular procedures of THA, aiming at the acetabular center (Fig. 1D). The final cup size was determined according the stability of the cup maintaining by the clamping force of the anterior and posterior aetabular walls. The bone defect above the cup was filled with bone wax or cement(Fig. 1E). The pelvis model was performed CT scan to obtain the data of the cup and bone defect (Fig. 1F). The model of the integral acetabular prosthesis was designed using 3-Matic 13.0 software (Materialise, Belgium) (Fig.1G and H).Four screws (red) pointed to the pubis, ischium, and posterosuperior part of the ilium, two screws (green) pointed to the sacroiliac joint (Fig. 1I). The blue area was designed using a 3D printed microporous structure on the bone-prosthesis interface to promote bone in-growth and long-term prosthetic stability. Ti6Al4 V was chosen as the material for the prosthesis (yellow part) (Fig. 1I). The integral acetabular prosthesis was manufactured using the Arcam A1 3D print machine (Arcam Corporation, Beijing, China) by means of the electron beam melting 3D printing technique (Fig. 1J). The integral customized acetabular prothesis was inserted into the acetabular socket with well press-fit(Fig. 1 K).The hip rotation center and bone defect were reconstructed anatomically(Fig. 1 L).

**Figure 1.**
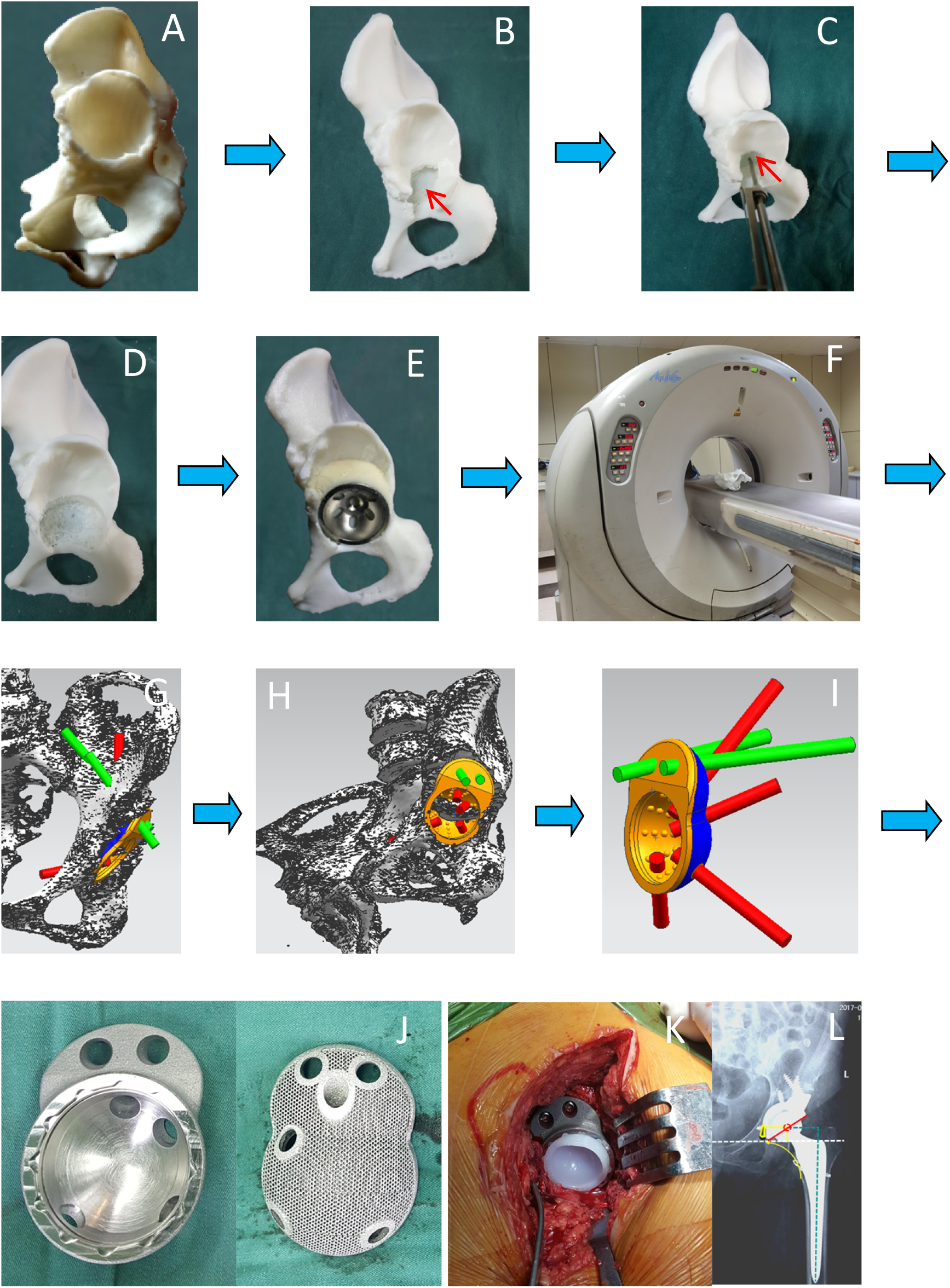
The flowchart of developing 3D printing integral customized acetabular prosthesis by model method. (A) :3D printing model. (B):Restoring Harris fossa. (C):Locating the acetabular center by ACL.(D):Reaming the acetabulum.(E):Installing the acetabular cup and filling the bone defect with bone wax/cement.(F):CT scan to obtain the data of the cup and bone defect.(G-I):Designing the integral customized acetabular prothesis.(J):3D printing integral customized acetabular prosthesis.(K): Installing integral customized acetabular cup in THA.(L):Postoperative X-ray. 3D=three-dimensional; ACL=acetabular center locator; CT=computer tomography;THA=total hip arthroplasty.

During the development process, we firstly needed to print the pelvis model using polylactic acid (PLA) material, secondly perform the simulated operation on the model, and then perform CT scan for the model to obtain the data of integral customized acetabular prosthesis. This process was time-consuming and laborious, and it was difficult to control the inclination and anteversion of the prosthesis accurately. Therefore, in this study, a novel method was developed to design the integral customized anatomical acetabular prosthesis on the basis of digital softwares, which could improve the efficiency and accuracy of prosthetic design. The purpose of this study was: (i) to introduce a new method to design the integral customized anatomical acetabular prosthesis by Mimics and 3-Matic softwares; (ii) to evaluate and compare the related indexes of prosthetic design between our previous model method and the novel software method.

## Methods

### Data sources

This study involved 15 patients (18 hips) who underwent primary cementless THA in the Department of Orthopedics of the First Affiliated Hospital of Bengbu Medical College from January 2015 to November 2021. The 15 patients included 3 men (3 hips) and 12 women (15 hips). The average age is 53.89 ± 8.48 years (range from 32 to 69 years). There were twelve hips of Crowe type II, six hips of Crowe type III.Inclusion criteria: ① age ≥18 years old ; ② Crowe type II/III DDH; ③ Tonis stage III hip osteoarthritis; ④ Consent to participate and cooperate with the investigators. Exclusion criteria: ① age <18 years old; ② Patients with normal developmental hip or Crowe type I and IV DDH; ③ Tonis stage 0-II hip osteoarthritis; ④ Tumors involving the acetabulum.

### Designing the integral customized anatomical acetabular prosthesis by Mimics and 3-Matic softwares (Software method)

Three dimensional (3D) pelvis CT scan (Brilliance 64-slice spiral CT, Philips Investments LTD., The Netherlands) was performed in all patients, and the data were imported into Mimics 10.01 (Materialise, Belgium) software. After removing bilateral femurs, the 3D pelvis mask was reconstructed. The shape of acetabular fossa was restored by removing the osteophytes using Edit Masks function(Fig. 2A). The data of 3D pelvis model was imported into 3-Matic software in STL format. The acetabular center was located at mean 28.7mm point (depending on acetabulum size) above the vertical bisection of the line between the anterior and posterior acetabular notches and the horizontal and vertical distance of hip center were mesured by Measure Distance function(Fig. 2B).The Design function was used to make a proper size hemisphere, the center of which pointed to the acetabular center vertically. The hollow function was used to make the hemisphere to be acetabular cup(Fig. 2C). The Align function was used to place acetabular cup into the acetabulum, the bottom of the cup reached the cortical bone of the acetabular fossa. The Measure Angle function was use to set the anteversion and inclination of the acetabular cup as 15 ° and 40 ° as accurate as possible(Fig. 2D and E). A solid ball with a radius larger than the acetabular cup was made to fill the superior bone defect. The solid ball was cut twice using the pelvis and acetabular cup as the cutting tool by the Design-Cut function. Hid and deleted extra parts. The Design-Cut function and Finish-Push function were used to reshape and polish the the wing of the acetabular prosthesis(Fig. 2G). The screws were designed as analytic cylinders with proper radius pointing to the pubis, ischium, and ilium respectively(Fig. 2H). The volume and surface area of the prosthesis’ wing were calculated(Fig. 2F).

**Figure 2.**
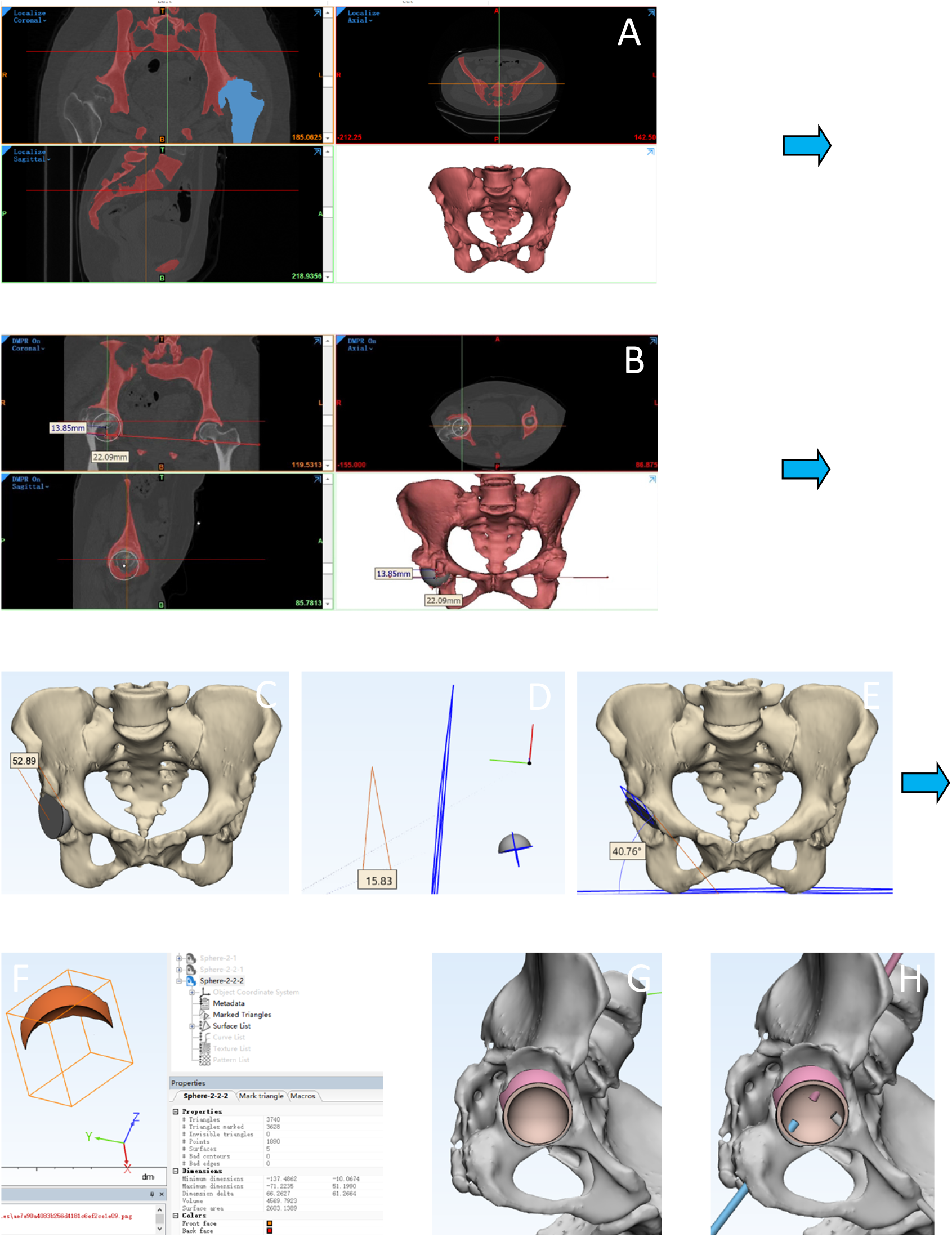
The research process of developing 3D printing integral customized acetabular prosthesis by the software method. (A) :Reconstructing the pelvis model by Mimics software.(B):Restoring hip center and measuring the horizontal and vertical distance by Mimics software. (C-E): Setting the anteversion and inclination of acetabular prothesis by 3-Matic software. (F):Measuring the volum and surface area of the wing part of acetabular prosthesis(bone defect) by 3-Matic software.(G,H):Installing the integral customized acetabular prothesis and setting the directions of scews pointing to the pubis, ischium, and ilium respectively by 3-Matic software.

### Designing the integral customized anatomical acetabular prosthesis by simulating operation on 3D printing model (Model method)

The pelvis model was printed by a 3D printing machine (Arigin 3DM400, Shanghai Arigin Medical Technology Co. LTD, China) using PLA material (Fig. 3A). The morphology of acetabular fossa was presented and restored by removing the superficial osteophytes (Fig. 3B). The acetabular center was located using acetabular center locator we developed. The acetabular socket was reamed by regular THA techniques, aiming at the acetabular center. Reaming was performed at a concentric circle of the acetabular center and to the depth of the acetabular fossa’s bottom. The final cup size was determined according the stability of the cup maintaining by the clamping force of the anterior and posterior acetabular walls (Fig. 3C). The bone defect above the cup was filled with bone wax (Fig. 3D). The volume of bone wax was calculated by a graduated cylinder using the drainage method(Fig. 3E,F).The area of bone defect was measured by the Photoshop method(Fig. 3G)(6). The surface of bone wax which contacted with the cup was covered with gauze precisely. The gauze was removed from the bone wax and laid flat. The gauze was placed in the area surrounded by two rulers which were mutually perpendicular. A picture was taken by digital camera (SONY DSC-T90, 10.2 MP, SONY Company, Tokyo, Japan). The pictures were inputted into Photoshop software. The boundary of gauze and rulers were marked respectively. The size of the selected zone could be expressed by the pixels. The actual area of gauze could be calculated by this method: the pixels of the gauze zone divided the pixels of the ruler zone and then multiplied the actual area of the ruler zone,such as (434*147) divided by (737*733) multiplied by (10*10) cm2. Plain films of the pelvis model were taken by C-arm X-ray machine(Siemens AG,Germany), and the images were imported into GeoGebraGeometry software (V6.0.574.0, America). The inclination was calculated by the classical method, that is, the angle between the elliptic (formed by the acetabular cup edge) long axis and the line of bilateral teardrops. The Lewinneck method was used to calculate the anteversion, that is, the anteversion was equal to arcsin (A/B). A and B were the short axis and the long axis of the ellipse formed by the acetabular cup edge. The vertical and horizontal distance of the hip rotation center were measured on the pelvic radiograph.

**Figure 3.**
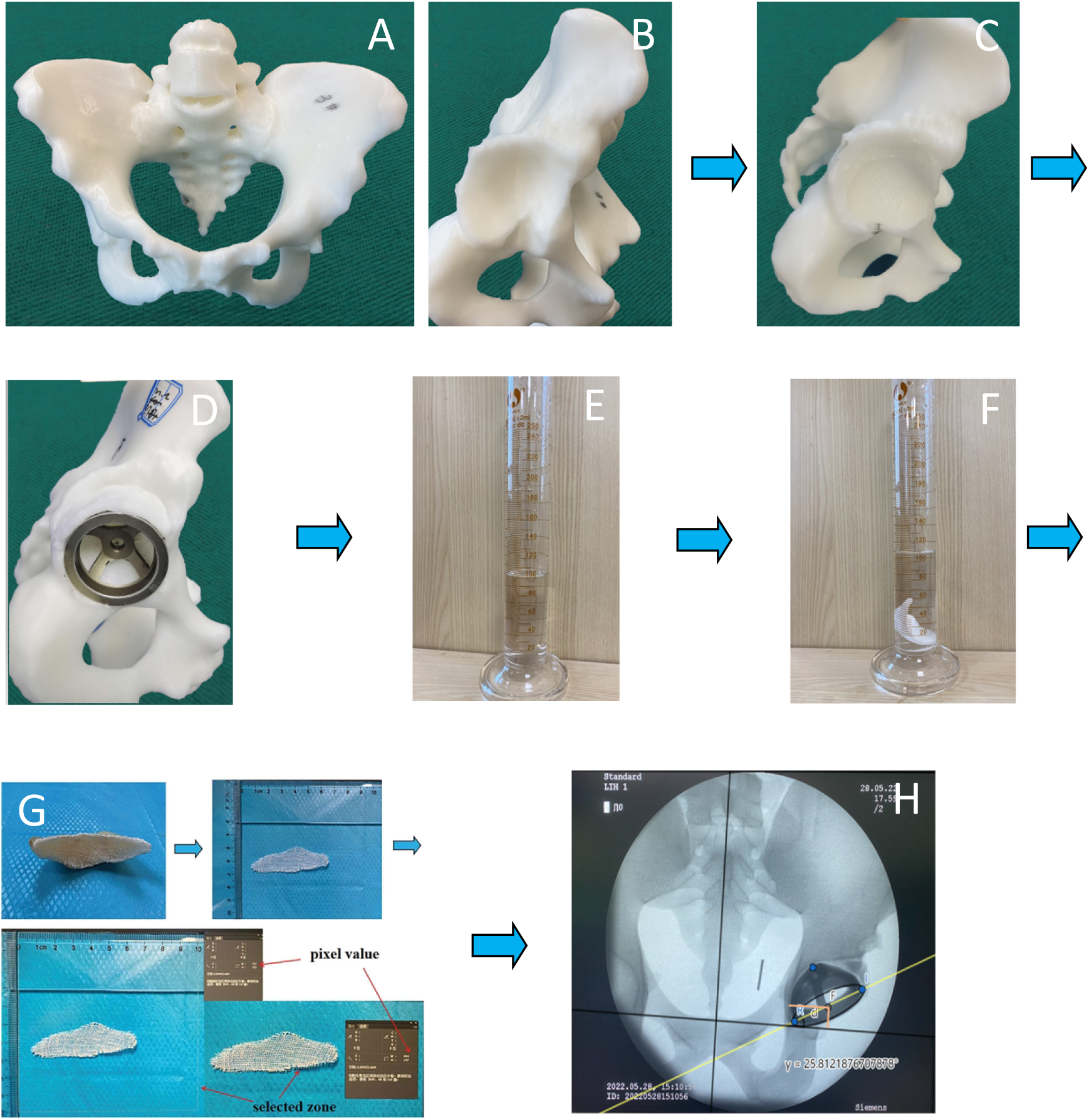
The research process of developing 3D printing integral customized acetabular prosthesis by the model method. (A) : 3D printing the pelvis model using PLA material. (B): the shell-like acetabulum and triangle acetabular fossa. (C) : Reaming the acetabulum.(D):Installing the acetabular test cup and filling the bone defect with bone wax. (E-F): Measuring the volum of bone defect. (G):Measuring surface area of bone defect.(H): Measuring the horizontal and vertical distance of hip center and the anteversion and inclination of acetabular cup on pelvis X-ray. PLA= Polylactic acid.

### Statistical Analysis

All statistical analyses were performed by using SPSS software for Windows (version 19.0; SPSS, Chicago, IL, USA). Continuous variables were presented as means and ranges. The 2-sided paired T test was used for comparison of cup size, hip rotation center data, the superficial area and volume of bone defect, the inclination and anteversion of the acetabular cup, and working time between two methods. A P value <0.05 was considered statistically significant.

## Results

The mean anteversion and inclination of acetabular cup in the software group and model group were(15.17±0.52)°and(40.24± 0.58)°, while the mean anteversion and inclination of acetabular cup in model group were (33.79 ± 13.43) °and (30.50 ± 11.03) °, the difference was statistically significant (t = 5.859, 3.767, P < 0.05). There were statistically significant difference between the software group and model group on the volume and superficial area of bone defect (t = 2.397, 2.707, P < 0.05). The mean working time in the software group was (0.47 ± 0.12) hours and (24.43 ± 0.68) in the model group, the difference was statistically significant (t = 138.51, P < 0.05).

There were no statistically significant difference between the software group and model group on cup size, the horizontal and vertical distance of hip center(t =1.458,0.114, 1.712, P > 0.05).

## Discussion

In total hip arthroplasty for patients with Crowe type II and III DDH, the application of novel 3D printed integral customized anatomical acetabular prosthesis could anatomically reconstruct the acetabulum and accurately repair bone defects, which was not only beneficial to restore the biomechanics of the hip joint, but also helped to realize equal length of lower limbs and improved the function of the gluteal medius. In addition, the complications of bone resorption and collapse of structural bone graft could be avoided. The clinical and radiological results of preliminary follow-up were satisfactory(12). In this study, two methods were introduced to design and develope of novel 3D printed integral customized anatomical acetabular prosthesis. The novel method based on digital softwares showed the advantages of more accurate prothesis position parameters, higher efficiency and lower design cost,compares with the model method(Table 1).

**Table 1.** The comparison of measurement results between the model method and software method 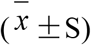 (n=18)

|  | Software method group | Model method group | t | P |
| --- | --- | --- | --- | --- |
| Cup size (mm) | $48.77 \pm 1.22$ | $48.56 \pm 1.15$ | 1.46 | 0.16 |
| Volume of bone defect (cm<sup>3</sup>) | $4.01 \pm 2.56$ | $5.06 \pm 2.86$ | 2.71 | 0.02 |
| Superficial area of bone defect (cm<sup>2</sup>) | $6.83 \pm 2.71$ | $8.31 \pm 2.21$ | 2.40 | 0.03 |
| Anteversion (°) | $15.17 \pm 0.52$ | $23.79 \pm 6.31$ | 5.86 | 0.00 |
| Inclination (°) | $40.24 \pm 0.58$ | $30.49 \pm 11.03$ | 3.77 | 0.002 |
| The horizontal distance of hip rotation center (mm) | $32.17 \pm 2.40$ | $32.08 \pm 1.80$ | 0.11 | 0.91 |
| The vertical distance of hip rotation center(mm) | $15.11 \pm 1.45$ | $14.36 \pm 1.53$ | 1.71 | 0.11 |
| Working time(h) | $0.47 \pm 0.12$ | $24.43 \pm 0.68$ | 138.51 | 0.00 |

It has been increasingly accepted and advocated that the hip rotation center should be restored anatomically in patients with Crowe type II/III DDH during THA(6, 10, 11). However, there is still lack of a simple, intuitive and operable method to reconstruct the hip rotation center and bone defect during the operation currently(13). Therefore, we developed a novel 3D printed integral customized anatomical acetabular prosthesis. The hip ration center was restored anatomically and there were no statistically significant difference between the software group and the model group on the restoration of hip rotation center. Other than some 2-dimensional methods, such as the template method, Ranawat method, Pierchon method,etc(14-16),we locate the acetabular center on average of 28.7mm (range, 25–31mm, depending on acetabulum size) above the vertical bisection of the line between the anterior and posterior acetabular notches as we have reported previously(3) in both groups. The installation angles of the acetabular cup play an important role in the life expectancy and postoperative stability of the prosthesis. Inappropriate anteversion and inclination are important factors affecting postoperative instability and dislocation of hip joint(17, 18). The difference between two groups on the anteversion and inclination of acetabular cup was statistically significant. It was difficult to control the anteversion and inclination during the reaming process on the pelvis model due to the complicated acetabular deformity in Crowe type II/III DDH, even for experienced surgeons.Gromov et al. (19) reported that it was easy to obtain excessive inclination and anteversion of the acetabular cup in DDH total hip arthroplasty due to the pursuit of the coverage and the initial stability of acetabular prosthesis. Zhao et al. (20) reported that the preoperative plan was in low accordance with the actual operation in acetabular cup installation position for patients with Crowe type II and III DDH due to the following reasons:the first reason was that the severe abnormal acetabular bone structures and the surrounding contracture soft tissues led to the difficulty in identifying anatomical reference landmarks to control the installation angle of the acetabular cup. Furthermore, the traditional free-hand technique of acetabular reaming was easy to deviate from the preoperative plan, especially in the anteversion, the reaming depth and the size of the acetabular cup.

Currently, the main solutions of Crowe type II and III DDH acetabular bone defect in THA include: granule bone graft, structural bone graft, tantalum metal augment, as well as bone cement(14, 21-23). Impaction granule bone graft is a reliable technique, which has high requirements for the surgeon to perform layer impaction(24). Structural bone graft is widely accepted by most orthopedists due to good initial stability for the acetabular cup and easy manipulation, however, it can not be avoided that high risks of the failure of osseointegration and subsequent bone resorption and collapse as well as prosthesis loosening(14). Tantalum metal augment can be used to reconstruct bone defects, which is believed to simplify the operation, but it is diffificult to match the shape of bone accurately(22). Bone cement method is a traditional technique, which is only used in elder patients with severe osteoporosis, due to high revision difficulty(23). Based on the preoperative three dimentional CT of the acetabulum, a novel 3D printing customized acetabular prothesis was developed to reconstruct the bone defect accurately with a perfect match. The initial stability could be obtained by the good press fit and individualized designed screws.There was no statistically significant difference between the software group and model group on cup size. The difference between two groups was statistically significant on the volume and surface area of bone defect. Both the software method and model method could restore the rotation center to the anatomical position, where the bone mass of acetabular anterior and posterior wall could be utilized to the utmost to determine the cup size. However, the difference between two groups on the mean anteversion and inclination of acetabular cup was statistically significant, which contributed to the difference of volume and surface area of bone defect. This significant difference in working time between two groups attributed to the complicated developing procedures of the model method and high efficiency of digital softwares.

Our study had some limitations. First, the sample size was too small to represent all cases of Crowe type II and III DDH. Second, the new methods were created to measure the horizontal and vertical distance of hip rotation center in the software group and the superficial area of bone defect in the model group, which need to be verified in the future study.Finally, the efficiency of two methods also needs to be further compared and investigated in clinical practice.

## Conclusions

In this work, a novel 3D printed integral customized acetabular prosthesis was developed by the model method and software method separately, which provides a promising way to restore the hip rotation center and reconstruct the bone defect in patients with Crowe type II and III DDH in THA. The results of this study indicated that the application of digital softwares could design and develope the novel 3D printed integral customized anatomical acetabular prosthesis in THA for DDH crowe type II and III with high accuracy and efficiency.

## Data Availability

The data underlying the results presented in the study

## Author contributions

Heng Zhang conceived the ideas, performed the experiment and drafted the manuscript. Jianning Zhao and Jiansheng Zhou revised the manuscript, reviewed the simulation results, and advised the organization of the main contents. Xiaodong Ma,Zengjing Cheng and Xuanxuan Li collected the detailed research results.

## Ethical Statement

This study was approved by the Ethics Committee on Human Research of the First Affiliated Hospital of Bengbu Medical College(2011053), and informed written consent was obtained from all involved patients.

